# Improvement in myocardial deformation but not structural indices in adults born preterm following a prescribed aerobic exercise intervention: a randomized clinical trial

**DOI:** 10.1101/2024.10.28.24316322

**Authors:** Winok Lapidaire, Afifah Mohamed, Wilby Williamson, Odaro J. Huckstep, Maryam Alsharqi, Cheryl M.J. Tan, Samuel Burden, Cameron Dockerill, William Woodward, Annabelle McCourt, Holger Burchert, Yvonne Kenworthy, Luca Biasiolli, Helen Dawes, Charlie Foster, Paul Leeson, Adam J. Lewandowski

## Abstract

**Background:** People born preterm (<37 weeks’ gestation) have a potentially adverse cardiac phenotype that progresses with blood pressure elevation. We sought to determine whether preterm-born adults with elevated and stage 1 hypertension exhibit similar cardiac structural and functional remodeling following a 16-week aerobic exercise intervention as their term-born peers.

**Methods:** We conducted an open, parallel, two-arm superiority randomized controlled (1:1) trial in n=203 adults aged 18-35 years old with elevated or stage 1 hypertension. Participants were randomized to a 16-week aerobic exercise intervention targeting three, 60-minute supervised sessions per week at 60-80% peak heart rate (exercise intervention group) or sign-posted to educational materials on hypertension and recommended lifestyle behaviors (control group). In a pre-specified cardiovascular magnetic resonance imaging (CMR) sub-study, CMR was performed on a 3.0-Tesla Siemens scanner to assess left ventricular (LV) and right ventricular (RV) structure and function before and after the 16-week intervention period. Group differences in outcome variables after the intervention were examined using analysis of covariance (ANCOVA) adjusting for baseline outcome variables, sex, and age. Interactions between birth category and intervention effect were investigated by including an interaction term in the intervention ANCOVA analyses.

**Results:** One hundred participants completed CMR scans both at baseline and after the 16-week intervention, with n=47 in the exercise intervention group (n=26 term-born; n=21 preterm-born) and n=53 controls (n=32 term-born; n=21 preterm-born). There was a significantly different response to the exercise intervention between preterm- and term-born participants. In term-born participants, LV mass to end-diastolic volume ratio decreased (adjusted mean difference: -3.43, 95% CI: -6.29,-0.56, interaction term p=0.027) and RV stroke volume index increased (adjusted mean difference: 5.53mL/m^2^, 95% CI: 2.60,8.47, interaction term p=0.076) for those in the exercise intervention group versus controls. No significant effects were observed for cardiac structural indices in preterm-born participants. In preterm-born participants, LV basal- and mid-ventricular circumferential strain increased (adjusted mean difference: -1.33, 95% CI: -2.07,-0.60, interaction term p=0.057 and adjusted mean difference: -1.54, 95% CI: -2.46,-0.63, interaction term p=0.046, respectively) and RV global longitudinal strain increased (adjusted mean difference=-1.99%, 95% CI=-3.12,-0.87, interaction p=0.053) for those in the exercise intervention group versus controls. No significant effects were observed for myocardial deformation parameters in term-born participants.

**Conclusions:** Aerobic exercise training induces improved myocardial function but not cardiac structure in preterm-born adults.

## INTRODUCTION

Preterm birth (<37 weeks’ gestation) affects more than 10% of births worldwide.^1^ Individuals born preterm are at greater risk of developing hypertension,^2^ new-onset heart failure,^3^ ischemic heart disease,^4^ and early cardiovascular-related mortality.^5^ Young adults born preterm have been observed to have a potentially adverse cardiac phenotype that progresses with blood pressure elevation, which may be relevant to their increased risk of cardiovascular disease.^6^ Compared to their term-born peers, preterm-born young adults have higher left ventricular (LV) and right ventricular (RV) mass indices alongside smaller internal LV and RV dimensions, with resulting lower LV and RV end-diastolic volumes.^7^ Each 1-mmHg elevation in systolic blood pressure (SBP) in adults born preterm associates with around double the increase in LV mass index and LV mass to end-diastolic volume ratio compared to term-born adults, suggesting a potentially greater cardiac vulnerability to blood pressure elevation.^6^

Young adulthood could be an optimal period to intervene in preterm-born individuals given the potential benefits of blood pressure control on cardiac remodeling decreases with older age.^8,9^ Although there is insufficient evidence to support the recommendation of starting antihypertensive medications in young adults with hypertension,^10^ aerobic exercise is known to positively impact cardiac structure and function with limited side effects.^11^ However, its effect in younger individuals with hypertension remains under-investigated^12^ and it is unknown whether either lifestyle or pharmacological interventions lead to beneficial LV and RV structural and/or functional improvement in preterm-born adults.

The Trial of Exercise to Prevent HypeRtension in young Adults (TEPHRA) is a randomized controlled exercise intervention trial in young adults with elevated blood pressure with a pre-specified sub-group of preterm-born participants and pre-specified cardiovascular magnetic resonance imaging (CMR) sub-study.^13^ The objective of the CMR sub-study was to determine whether preterm-born adults with elevated and stage 1 hypertension, who had no history of antihypertensive medication use, exhibit similar LV and RV remodeling following a 16-week aerobic exercise intervention as their term-born peers with comparable blood pressures.

## METHODS

### Study design

We conducted an open, parallel, two-arm superiority randomized controlled (1:1) trial in n=203 participants as previously described.^13,14^ Briefly, participants completed a baseline study visit and were then randomized to a 16-week aerobic exercise intervention targeting three, 60-minute supervised sessions per week at 60-80% peak heart rate (exercise intervention) or sign-posted to educational materials (controls). Participants completed a follow-up study visit after the 16-week intervention period that was identical to the baseline study visit.

Randomization was performed using a computerized randomization program (Sealed Envelope^TM^; https://www.sealedenvelope.com/). A minimization algorithm (with a random element of 80%) was used to ensure balanced allocation across the two groups for key prognostic factors: sex (male/female); age (<24 years old, 24– 29 years old, 30–35 years); and gestational age of participants (≤32 weeks, 32–37 weeks, >37 weeks). In a pre-specified CMR sub-study in n=100 individuals, CMR was performed before and after the 16-week intervention.^13^ The pre-specified recruitment strategy was designed to recruit a higher percentage of preterm-born participants than in the general population (Figure 1). The outcome assessors were not involved in randomization or intervention delivery and remained blinded until after completion of final analysis.

**Figure 1:**
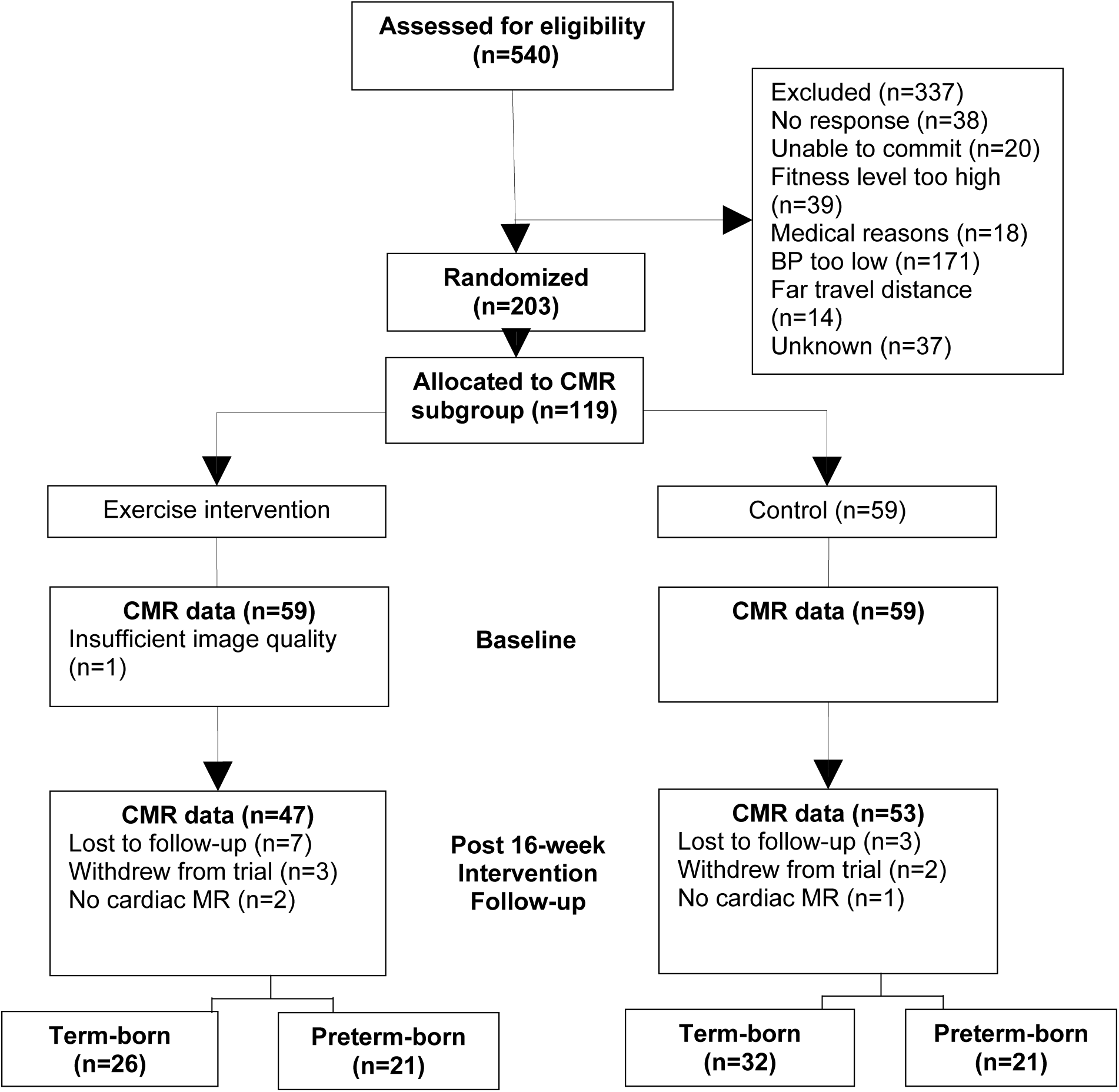
Trial profile for the cardiovascular magnetic resonance imaging (CMR) sub-study.

### Study population

Participants were recruited through open recruitment; general practitioner records; invitations from hospital birth registers; online advertising on Facebook, Instagram, and Twitter; and invitations after participation in previous studies. Participants were aged 18-35 years with: elevated or stage 1 hypertension (24-hour awake ambulatory SBP and/or diastolic blood pressure (DBP) >115/75 mmHg, but <159/99 mmHg); a body mass index (BMI) <35kg/m^2^; not on, and had not previously been prescribed, antihypertensive medications; had a verifiable birth history of preterm birth (<37 weeks) or full-term birth (≥37 weeks); and had the ability to access and use a computer and the internet. Exclusion criteria were participation in structured exercise greater than once per week or with high self-reported cardiovascular fitness; contraindications to exercise; unable to walk briskly on the flat for 15 minutes; and any evidence of cardiomyopathy, inherited cardiac abnormalities or other significant cardiovascular disease. All preterm-born participants from the main TEPHRA study were invited to join the CMR sub-study, while term-born participants of the main study were invited until n=100 participants were recruited for the CMR sub-study. Enrollment occurred between June 30, 2016 and October 26, 2018; the final follow-up was completed on January 9, 2020.

The trial protocol and any subsequent amendments were approved by the University of Oxford as host institution and study sponsor and by the South Central Research Ethics Committee for the National Health Service Health Research Authority (Reference: 16/SC/0016). The trial was registered on Clinicaltrials.gov on March 30, 2016 (NCT02723552). A trial steering committee and independent data and safety monitoring board monitored the study, and all participants provided written informed consent. The investigators ensured that the study was conducted in accordance with the principles of the Declaration of Helsinki as well as in accordance with relevant regulations and good clinical practice.

### Intervention

The intervention stipulated three 60-minute aerobic training sessions (completed on bicycle ergometers) on separate days per week for 16 weeks at an exercise intensity of 60–80% of peak heart rate measured at baseline by cardiopulmonary exercise testing. A wrist-worn heart rate and activity monitor (Fitbit Charge HR; Fitbit, Inc.) was gifted to the participants who were encouraged to wear it daily. To track physical activity in the intervention group, activity from the wrist-worn activity monitor was tracked using the Fitabase data management platform and records kept of training sessions attended. The compliance threshold for the intervention was set at 80%, equivalent to 39 or more independent training sessions, with no more than two weeks between sessions. A compliant session was defined a priori as a supervised one. This definition was refined by the trial committees during the trial to being an aerobic session defined as a supervised gym session 40 to 60 minutes long; a self-reported aerobic session 40 to 60 minutes long; a day with a step count of 8,000 steps or more measured by their Fitbit; or a total of 40 or more Fitbit active minutes that were defined as fairly and vigorously active. The intervention team consisted of physiologists, physiotherapists, clinical nurse specialists, and a physician.^13,14^

### Study visit

The full trial protocol has previously been published.^13^ Study visits took place at the Oxford Cardiovascular Clinical Research Facility and Oxford Centre for Clinical Magnetic Resonance Research at the John Radcliffe Hospital, Oxford, UK. Blood pressure was measured using the automated mode of a validated sphygmomanometer (Dinamap V100, GE Healthcare) after five minutes of seated rest. Three seated blood pressure measurements were done once every minute, with the second and third blood pressure readings averaged for analyses. Height and weight were measured to the nearest centimeter and 0.1 kg, respectively, with participants’ footwear removed and light clothing worn.

CMR was performed on a 3.0-Tesla Siemens TIM Trio scanner (Siemens Healthineers) to assess cardiac and aortic structure and function. Cardiac horizontal and vertical long-axis and LV outflow tract retrospective electrocardiography (ECG) gated steady-state free precession (SSFP) cine images were acquired, followed by cardiac short-axis SSFP cine images. All cardiac cine images were acquired during end-expiratory breath-hold. Short-axis cine images were obtained from the base to apex of the heart. Aortic ECG-gated, SSFP transverse cine images were acquired during end-expiratory breath-hold at the level of the right pulmonary artery showing cross-sections of the ascending aorta (AA) and proximal descending aorta (PDA). Blood pressure was measured immediately following the aortic cine image acquisition using the Vicorder system (SMT Medical) to estimate central blood pressure.^15^ Further details on the CMR protocol can be found in the main protocol paper.^13^

### CMR Image Analysis

Cardiac analysis was done using analytic software (CVI42; Circle Cardiovascular Imaging). To evaluate LV and RV volumes, mass, and dimensions, both the epicardial and endocardial borders were manually contoured on short-axis cine images for each slice at end diastole and endocardial borders at end systole. The end-diastolic and end-systolic cardiac phases and basal and apical LV and RV slices were visually determined as previously described.^6,16^ LV and RV volumes and mass measurements were indexed to body surface area. Length of the LV was measured as the length from the LV apex to the middle of the mitral valve annulus at end diastole.

Myocardial deformation analysis was performed using feature tracking. Endocardial and epicardial borders of the LV and RV for the horizontal long-axis cine, and the LV for the basal, mid, and apical short-axis cines, were manually contoured on the end-diastolic frame. The deformation of the myocardium was then automatically tracked through the phases of the cardiac cycle. Aortic distensibility was calculated according to a fully automated image analysis workflow using Matlab software (Mathworks Inc).^17^ Briefly, maximum and minimum aortic cross-sectional areas over the cardiac cycle were determined by considering systolic-diastolic variation in cross-sectional lumen area of the AA and PDA. Aortic distensibility was calculated as the relative change in vessel areas divided by the pulse pressure.

### Statistical analysis

Statistical analyses were run in R (version 4.0.3). Continuous variables are presented as mean (standard deviation) when the data were normally distributed and median (interquartile range) when the data were non-normally distributed. Frequencies are presented as numbers with percentages. Group differences in outcome variables at follow-up between the exercise intervention group and control group were analyzed using analysis of covariance (ANCOVA), adjusting for baseline outcome variables and sex, age, and gestational age category (<32, 32-37, or >37 weeks), and are presented as adjusted means and confidence intervals (CI). Subgroups (term- and preterm-born) were defined by splitting the gestational age variable from the minimization procedure (gestation: <32, 32-37, or >37 weeks) into two levels: >37 and <37 weeks’ gestation. The subgroup analysis was then performed by fitting additional linear models, including an interaction term between treatment allocation (exercise, control) and gestational age category (<37 and >37 weeks) to test whether the intervention effects would vary significantly across subgroups. P values <0.05 and 95% CI were used to indicate statistical significance.

## RESULTS

### Baseline characteristics

One hundred participants completed CMR scans at both the baseline study visit and follow-up visit after the 16-week intervention, with n=47 in the exercise intervention group (n=26 term-born; n=21 preterm-born) and n=53 in the control group (n=32 term-born; n=21 preterm-born) (Figure 1). Baseline participant characteristics for the preterm-born and term-born participants in the exercise intervention and control arms of the trial can be found in Table 1. Age, sex, BMI, and blood pressure were all similar across the groups and trial arms.

**Table 1:** Participant characteristics of exercise intervention and control groups in the cardiovascular magnetic resonance sub-study, separated by preterm-born and term-born participants

|  | Exercise Intervention |  | Control |  |
| --- | --- | --- | --- | --- |
|  | Term<br>n=26 | Preterm<br>n=21 | Term<br>n=32 | Preterm<br>n=21 |
| Age, years | 28.7 (3.5) | 28.9 (4.8) | 27.2 (4.2) | 29.5 (4.6) |
| Male, n (%) | 14 (54) | 16 (50) | 11 (52) | 9 (43) |
| Employed, n (%) | 24 (96) | 30 (97) | 20 (100) | 19 (95) |
| University Degree, n (%) | 22 (88) | 27 (87) | 13 (65) | 16 (80) |
| Family history of cardiovascular disease, n (%) | 9 (35) | 10 (31) | 12 (57) | 9 (43) |
| BMI, kg/m<sup>2</sup> | 25.3 (3.9) | 24.2 (2.5) | 25.7 (2.8) | 23.7 (4.1) |
| Smokers, n (%) | 0 (0) | 3 (10) | 0 (0) | 2 (10) |
| Units of alcohol per week | 6.8 (5.4) | 5.0 (5.5) | 3.9 (5.0) | 7.0 (10.7) |
| Awake ambulatory blood pressure, mmHg |  |  |  |  |
| Systolic | 129.5 (11.4) | 129.6 (10.4) | 129.8 (8.9) | 127.8 (8.7) |
| Diastolic | 78.2 (6.4) | 76.5 (6.8) | 78.5 (7.3) | 77.0 (7.7) |
| Oxygen uptake peak, mL/kg/min | 34.0 (7.7) | 33.9 (6.7) | 35.8 (7.9) | 33.0 (6.0) |
| Cholesterol-HDL ratio | 3.5 (1.0) | 3.3 (1.0) | 3.6 (1.1) | 2.9 (1.3) |
| Glucose, mmol/L | 4.8 (0.5) | 4.9 (0.3) | 4.9 (0.4) | 4.7 (0.5) |
| HOMA insulin resistance | 1.0 (0.5) | 0.9 (0.4) | 1.0 (0.4) | 0.8 (0.3) |

*Values presented as mean (standard deviation) unless stated otherwise. BMI stands for body mass index; HDL, high density lipoprotein; HOMA, Homeostatic Model Assessment for Insulin Resistance.*

### Response to exercise intervention

The significant effects of the exercise intervention on cardiac structural measures, when preterm-born and term-born participants were grouped together, included increases in LV and RV ejection fraction, LV and RV stroke volume index, and LV length. Additionally, LV basal, mid, and apical circumferential strain, RV end-diastolic volume, and RV global longitudinal strain increased (Supplementary Tables 1 and 2).

### Unique cardiac phenotype in adults born preterm

At baseline, there were statistically significant differences in CMR measures between term-born and preterm-born participants in all structural and functional LV and RV parameters, except LV end-diastolic volume index (Table 2). The statistically significant differences, comparing preterm-born to term-born participants, included: lower LV and RV mass index, LV and RV stroke volume index, LV length, LV and RV ejection fraction, LV and RV global longitudinal strain, LV basal, mid, and apical circumferential strain, and RV end-diastolic volume index, as well as higher LV mass/end-diastolic volume. There were no significant differences between the preterm-born and term-born participants for aortic distensibility measures at baseline.

**Table 2:**
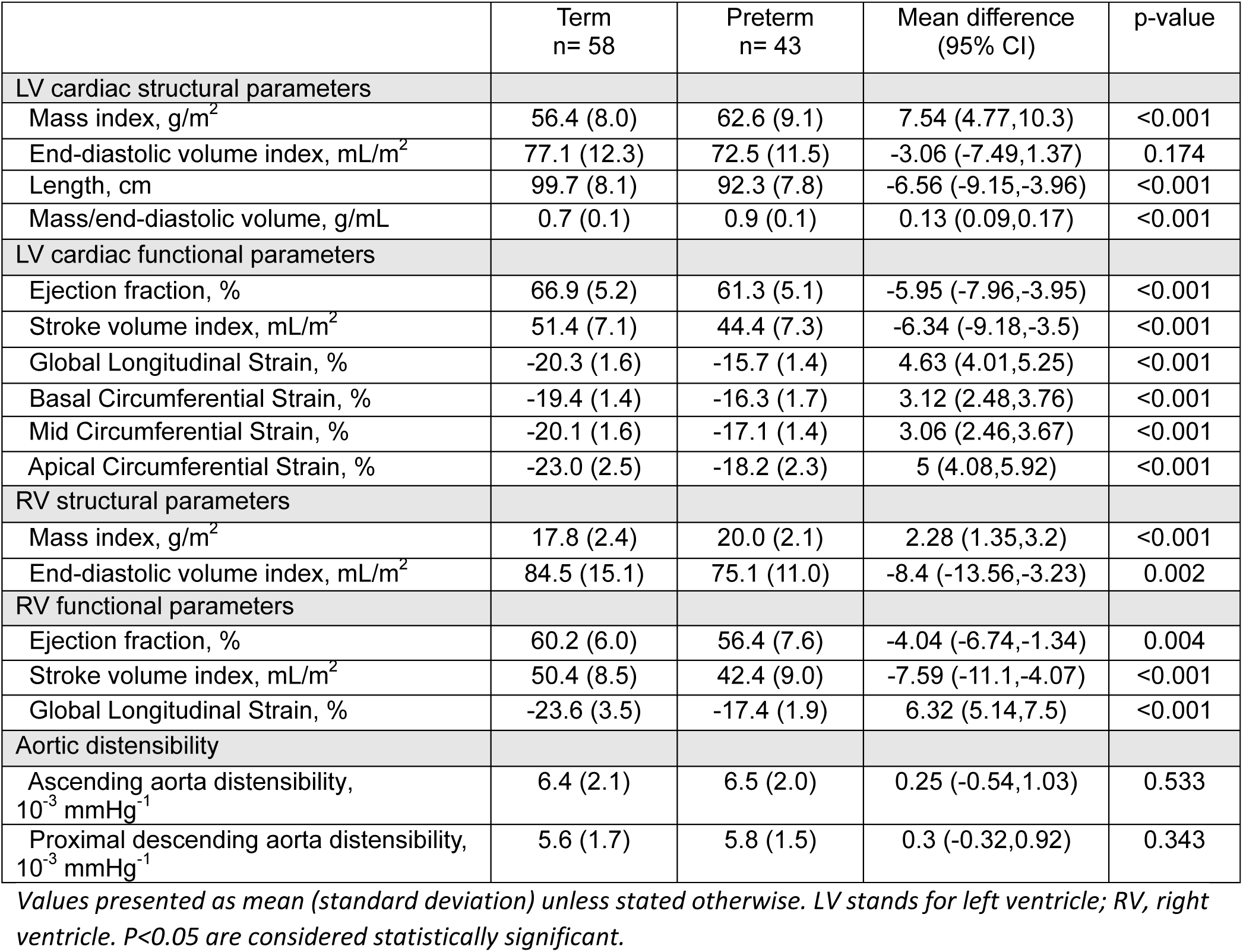
Cardiovascular magnetic resonance measures for preterm-born and term-born participants at baseline

|  | Term<br>n= 58 | Preterm<br>n= 43 | Mean difference<br>(95% CI) | p-value |
| --- | --- | --- | --- | --- |
| LV cardiac structural parameters |  |  |  |  |
| Mass index, g/m<sup>2</sup> | 56.4 (8.0) | 62.6 (9.1) | 7.54 (4.77,10.3) | <0.001 |
| End-diastolic volume index, mL/m<sup>2</sup> | 77.1 (12.3) | 72.5 (11.5) | -3.06 (-7.49,1.37) | 0.174 |
| Length, cm | 99.7 (8.1) | 92.3 (7.8) | -6.56 (-9.15,-3.96) | <0.001 |
| Mass/end-diastolic volume, g/mL | 0.7 (0.1) | 0.9 (0.1) | 0.13 (0.09,0.17) | <0.001 |
| LV cardiac functional parameters |  |  |  |  |
| Ejection fraction, % | 66.9 (5.2) | 61.3 (5.1) | -5.95 (-7.96,-3.95) | <0.001 |
| Stroke volume index, mL/m<sup>2</sup> | 51.4 (7.1) | 44.4 (7.3) | -6.34 (-9.18,-3.5) | <0.001 |
| Global Longitudinal Strain, % | -20.3 (1.6) | -15.7 (1.4) | 4.63 (4.01,5.25) | <0.001 |
| Basal Circumferential Strain, % | -19.4 (1.4) | -16.3 (1.7) | 3.12 (2.48,3.76) | <0.001 |
| Mid Circumferential Strain, % | -20.1 (1.6) | -17.1 (1.4) | 3.06 (2.46,3.67) | <0.001 |
| Apical Circumferential Strain, % | -23.0 (2.5) | -18.2 (2.3) | 5 (4.08,5.92) | <0.001 |
| RV structural parameters |  |  |  |  |
| Mass index, g/m<sup>2</sup> | 17.8 (2.4) | 20.0 (2.1) | 2.28 (1.35,3.2) | <0.001 |
| End-diastolic volume index, mL/m<sup>2</sup> | 84.5 (15.1) | 75.1 (11.0) | -8.4 (-13.56,-3.23) | 0.002 |
| RV functional parameters |  |  |  |  |
| Ejection fraction, % | 60.2 (6.0) | 56.4 (7.6) | -4.04 (-6.74,-1.34) | 0.004 |
| Stroke volume index, mL/m<sup>2</sup> | 50.4 (8.5) | 42.4 (9.0) | -7.59 (-11.1,-4.07) | <0.001 |
| Global Longitudinal Strain, % | -23.6 (3.5) | -17.4 (1.9) | 6.32 (5.14,7.5) | <0.001 |
| Aortic distensibility |  |  |  |  |
| Ascending aorta distensibility,<br>10<sup>-3</sup> mmHg<sup>-1</sup> | 6.4 (2.1) | 6.5 (2.0) | 0.25 (-0.54,1.03) | 0.533 |
| Proximal descending aorta distensibility,<br>10<sup>-3</sup> mmHg<sup>-1</sup> | 5.6 (1.7) | 5.8 (1.5) | 0.3 (-0.32,0.92) | 0.343 |

Values presented as mean (standard deviation) unless stated otherwise. LV stands for left ventricle; RV, right ventricle. P<0.05 are considered statistically significant.

### Differing cardiac structural remodeling in response to exercise intervention dependent on birth history

The exercise intervention effects differed between preterm-born and term-born participants for LV mass/end-diastolic volume (interaction p=0.027). Term-born participants in the exercise intervention group decreased LV mass to end-diastolic volume ratio (adjusted mean difference=-3.43, 95% CI=-6.29,-0.56), while preterm-born participants demonstrated an increase in this metric (Figure 2). Although RV mass index reduced in the preterm-born group (adjusted mean difference=-0.87g/m^2^, 95% CI=-1.86,-0.05), the interaction term did not reach statistical significance (p=0.19). Two preterm-born participants and one term-born participant in the exercise intervention arms did not meet the compliance threshold.

**Figure 2:**
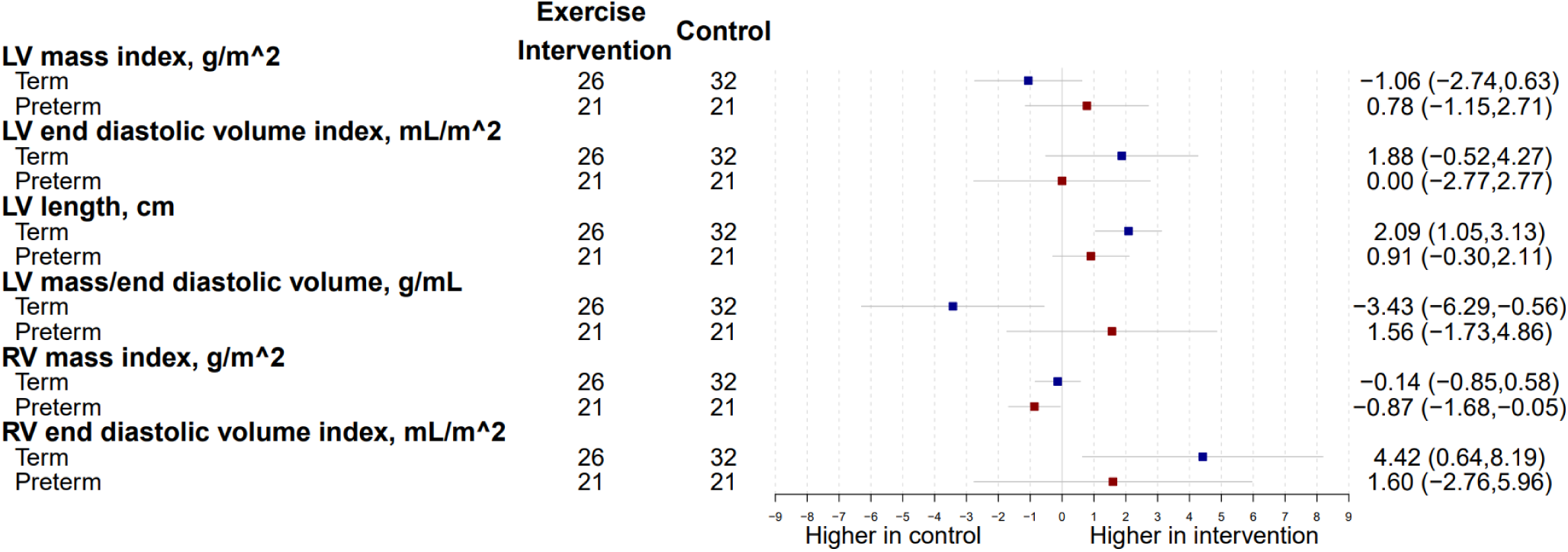
Differences between exercise intervention and control groups for preterm-born (red) and term-born (blue) participants for left ventricular (LV) and right ventricular (RV) structural parameters after the 16-week intervention. Change is presented as adjusted mean in the outcome of interest, in the units for that outcome, with 95% confidence interval error bars.

### Cardiac volumetric functional and myocardial deformation changes dependent on birth history

The cardiac volumetric functional parameters consistently demonstrated significant increases in the term-born participants but not preterm-born participants for the exercise intervention group following the 16-week intervention (Figure 3). In term-born participants, LV ejection fraction (adjusted mean difference=1.87%, 95% CI:0.10,3.64), interaction p=0.43), RV ejection fraction (adjusted mean difference=2.86%, 95% CI=0.52,5.20, interaction p=0.28), LV stroke volume index (adjusted mean difference=2.84 mL/m^2^, 95% CI=0.54,5.14, interaction p=0.14), RV stroke volume index (adjusted mean difference=5.53mL/m^2^, 95% CI=2.60,8.47, interaction p=0.076) all increased in the exercise intervention group, but the interaction effects were at trend level or did not reach statistical significance. Conversely, functional myocardial deformation changes in response to the exercise intervention only reached statistical significance in the preterm-born participants and not in the term-born participants (Figure 4). These changes included improved LV and RV global longitudinal strain (adjusted mean difference=-1.18, 95% CI=-2.07,-0.30, interaction p=0.10 and adjusted mean difference=-1.99%, 95% CI=-3.12,-0.87, interaction p=0.053), as well as improved LV basal, mid, and apical circumferential strain (adjusted mean difference=-1.33%, 95% CI=-2.07,-0.60, interaction p=0.057; adjusted mean difference=-1.54%, 95% CI: -2.46,-0.63, interaction p=0.046; adjusted mean difference=-1.04%, 95% CI=-2.06, -0.02, interaction p=0.41, respectively). However, some of the interaction effects were at trend level or did not reach statistical significance.

**Figure 3:**
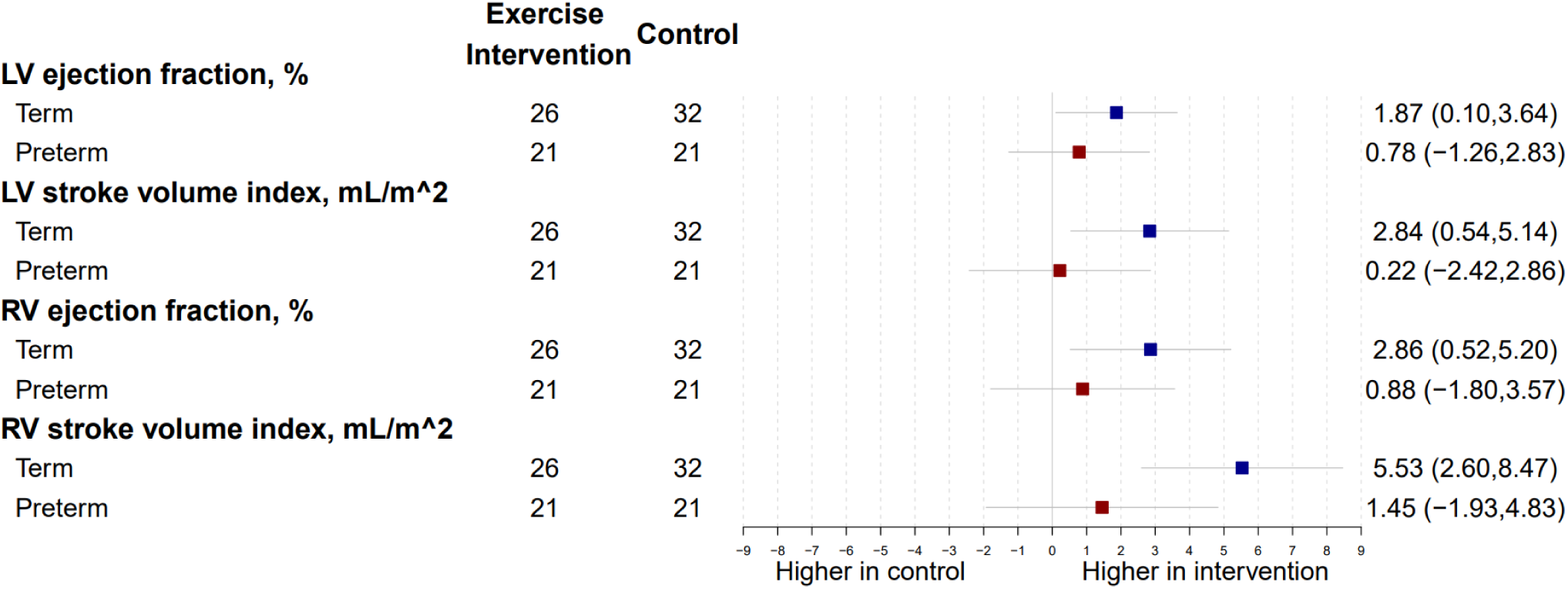
Differences between exercise intervention and control groups for preterm-born (red) and term-born (blue) participants for left ventricular (LV) and right ventricular (RV) functional parameters after the 16-week intervention. Change is presented as adjusted mean in the outcome of interest, in the units for that outcome, with 95% confidence interval error bars.

**Figure 4:**
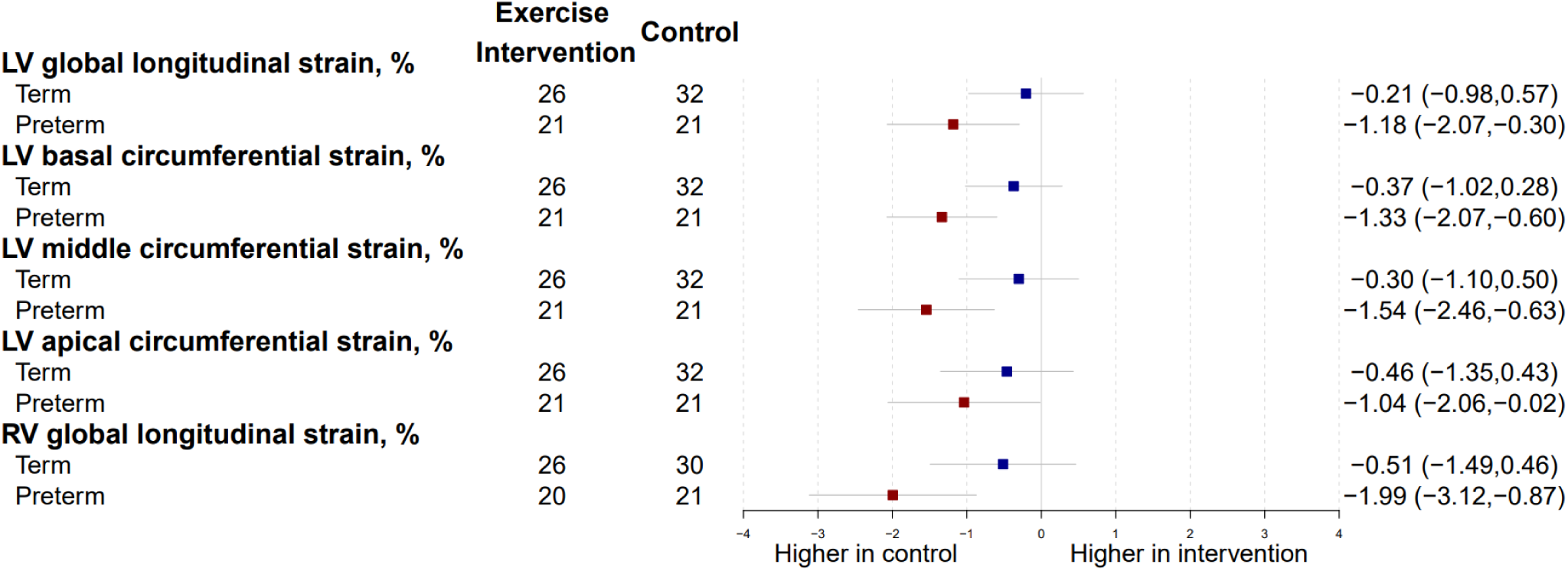
Differences between exercise intervention and control groups for preterm-born (red) and term-born (blue) participants for left ventricular (LV) and right ventricular (RV) myocardial deformation parameters after the 16-week intervention. Change is presented as adjusted mean in the outcome of interest, in the units for that outcome, with 95% confidence interval error bars.

### Blood pressure changes dependent on birth history

There were no significant interaction effects between preterm birth history and exercise intervention effect on systolic or diastolic blood pressure parameters (interaction p=0.46 and p=0.62, respectively). There was no statistically significant exercise intervention effect for systolic blood pressure in either the term-born participants (adjusted mean difference=1.40mmHg, 95% CI=-2.29,5.08) or preterm-born participants (adjusted mean difference=-0.71mmHg, 95% CI=-4.95,3.53). Diastolic blood pressure was also not significantly impacted by the exercise intervention in the term-born participants (adjusted mean difference=3.14mmHg, 95% CI=0.04,6.24) or preterm-born participants group (adjusted mean difference=1.95mmHg, 95% CI=-1.59,5.49).

## DISCUSSION

This trial demonstrates for the first time that young adults born preterm with elevated and stage 1 hypertension have an altered cardiac remodeling response to prescribed aerobic exercise training compared to their term-born peers. Prior to the intervention, preterm-born adults showed potential cardiac structural and functional impairments compared to term-born adults, including greater LV and RV mass, lower LV and RV ejection fractions and stroke volumes, as well as impaired LV and RV myocardial deformation parameters. After the 16-week intervention period, there were no changes observed for cardiac structural and volumetric-derived functional parameters in preterm-born adults, but term-born adults in the exercise intervention group decreased LV mass to end-diastolic volume ratio and increased RV stroke volume compared to participants in the control arm. Conversely, following the 16-week intervention period, there were no changes observed in myocardial deformation parameters in the term-born adults, but preterm-born adults in the exercise intervention group increased RV longitudinal strain and LV basal-and mid-ventricular circumferential strain compared to participants in the control arm. These results could not be explained by a differing response in blood pressure to the exercise intervention.

Underlying differences in cardiac phenotype may explain why preterm-born adults in our current trial did not exhibit improvements in LV and RV structural or functional volumetric parameters following the 16-week exercise intervention. Birth is characterized by significant changes in cardiac flow dynamics and cellular signaling pathways in the newborn baby that trigger a shift in cardiomyocyte energy utilization, contractility, and growth from a primarily hyperplastic to hypertrophic pattern.^18^ Experimental ovine and rodent models have demonstrated this shift also occurs at the time of preterm birth when the myocardium is relatively immature,^19,20^ resulting in pathological changes in cardiomyocyte volume and function that lead to increased risk of heart failure.^7,21,22^ This early, altered trajectory of cardiac development also appears in neonates and infants born preterm and appears to persist into adulthood.^7^ In a study by Mohamed et al. including 468 adults (200 born preterm; 268 born term) who underwent CMR imaging, it was observed that changes in LV mass and LV mass to end-diastolic volume ratio for each 1-mmHg increase in SBP were two-times greater in preterm-born adults compared with term-born adults.^6^ Importantly, for the same SBP, these cardiac measures are also higher in the preterm-born adults compared to their term-born peers, even after adjustment for potential confounders including age, sex, birthweight z-score, and BMI, supporting the hypothesis that the preterm birth related disruption in early cardiac development may track into adulthood.^23^ Indeed, it is plausible that some of these cardiac changes become fixed, or programmed, early in life and lead to greater cardiac vulnerability to secondary insults, such as hypertension.^24^

Observational studies have shown an impaired LV and RV response to acute exercise stress in adults born preterm. Specifically, in a study using echocardiography at rest and during exercise on an upright cycle ergometer, it was shown that LV ejection fraction and cardiac index in the preterm-born adults did not increase to the extent observed in term-born adults.^25^ Similarly, in a study by Goss et al. using right heart catherization at rest and during exercise on a supine ergometer, it was shown that RV cardiac index and stroke work did not increase in preterm-born adults to the extent observed in term-born adults.^26^ Despite these observed impairments in LV and RV volume reserve, preterm-born adults have been shown to have a greater myocardial contractile reserve under acute hypoxic stress conditions than their term-born peers.^27^ Our trial results support this greater myocardial contractile reserve given that preterm-born adults, despite lower LV and RV myocardial strain measures at baseline, showed a significant increase in both LV and RV myocardial strain measures in the exercise intervention group compared to controls, which was not observed in term-born adults.

There were limitations to this single center trial. Although the intervention period was relatively short, the duration was in line with other similar trials in young adults with hypertension^12^ and was sufficient to see a significant improvement in fitness in the form of cardiopulmonary exercise capacity^28^ and cardiac remodeling, especially in the term-born adults. Nevertheless, it is plausible that a longer intervention period, or alternative form of exercise such as high intensity interval training,^29^ may be needed to observe greater improvements in cardiac structure and function for both preterm- and term-born adults. While it was not possible to mask participants to trial group allocation, researchers conducting data collection study visits and performing statistical analyses were blinded to group allocation. Furthermore, group sizes for the CMR sub-study, once broken down by those born preterm and term, were relatively small. This is a likely explanation for why many of the interaction terms for the observed trial allocation group differences based on preterm and term birth status did not reach statistical significance. The sample size also meant the trial did not have sufficient power to explore to what extent the degree of prematurity, or other associated perinatal complications, may explain our results and will be an important consideration for designing future related trials.

## CONCLUSIONS

We have demonstrated in this exploratory sub-group analysis of individuals with elevated blood pressure and stage 1 hypertension that those born preterm have a differing cardiac remodeling response to aerobic exercise training than their term-born peers. Further work is needed to determine whether modified exercise protocols or combination with pharmacological intervention may lead to additional cardiac remodeling benefits.

## Data Availability

Data are available upon request.

